# Freezing of gait assessment with inertial measurement units and deep learning: effect of tasks, medication states, and stops

**DOI:** 10.1101/2023.05.05.23289387

**Authors:** Po-Kai Yang, Benjamin Filtjens, Pieter Ginis, Maaike Goris, Alice Nieuwboer, Moran Gilat, Peter Slaets, Bart Vanrumste

## Abstract

**Background:** Freezing of gait (FOG) is an episodic and highly disabling symptom of Parkinson’s Disease (PD). Traditionally, FOG assessment relies on time-consuming visual inspection of camera footage. Therefore, previous studies have proposed portable and automated solutions to annotate FOG. However, automated FOG assessment is challenging due to gait variability caused by medication effects and varying FOG-provoking tasks. Moreover, whether automated approaches can differentiate FOG from typical everyday movements, such as volitional stops, remains to be determined. To address these questions, we evaluated an automated FOG assessment model with deep learning (DL) based on inertial measurement units (IMUs). We assessed its performance trained on all standardized FOG-provoking tasks and medication states, as well as on specific tasks and medication states. Furthermore, we examined the effect of adding stopping periods on FOG detection performance.

**Methods:** Twelve PD patients with self-reported FOG (mean age 69.33 ± 6.28 years) completed a FOG-provoking protocol, including timed-up-and-go and 360-degree turning-in-place tasks in On/Off dopaminergic medication states with/without volitional stopping. IMUs were attached to the pelvis and both sides of the tibia and talus. A multi-stage temporal convolutional network was developed to detect FOG episodes. FOG severity was quantified by the percentage of time frozen (%TF) and the number of freezing episodes (#FOG). The agreement between the model-generated outcomes and the gold standard experts’ video annotation was assessed by the intra-class correlation coefficient (ICC).

**Results:** For FOG assessment in trials without stopping, the agreement of our model was strong (ICC(%TF) = 0.92 [0.68, 0.98]; ICC(#FOG) = 0.95 [0.72, 0.99]). Models trained on a specific FOG-provoking task could not generalize to unseen tasks, while models trained on a specific medication state could generalize to unseen states. For assessment in trials with stopping, the model trained on stopping trials made fewer false positives than the model trained without stopping (ICC(%TF) = 0.95 [0.73, 0.99]; ICC(#FOG) = 0.79 [0.46, 0.94]).

**Conclusion:** A DL model trained on IMU signals allows valid FOG assessment in trials with/without stops containing different medication states and FOG-provoking tasks. These results are encouraging and enable future work investigating automated FOG assessment during everyday life.

## Background

Parkinson’s disease (PD) is a neurodegenerative disorder that affects over six million people worldwide (1). One of the most debilitating gait disorders associated with PD is freezing of gait (FOG), which develops in approximately 70% of PD patients over the course of their disease (2; 3). Clinically, FOG is defined as a “brief, episodic absence or marked reduction of forward progression of the feet despite the intention to walk” and is often divided into three manifestations based on leg movement: 1) trembling: tremulous oscillations in the legs of 8-13 Hz; 2) shuffling: very short steps with poor clearance of the feet; and 3) complete akinesia: no visible movement in the lower limbs (1; 4). While one patient can experience different FOG manifestations, the distribution of manifestations can vary widely among individuals, in which trembling and shuffling are more common than akinetic freezing (5). The unpredictable nature of FOG poses a significant risk of falls and injuries for PD patients (6; 7; 8), and it can also affect their mental health and self-esteem, leading to a lower quality of life (9). To relieve the symptoms, dopaminergic medication such as Levodopa is mainly used (10). During Off-medication states, FOG more commonly occurs (11), while in contrast, FOG episodes are milder in On-medication states but may manifest differently with more trembling (12).

To qualitatively assess FOG severity in PD patients and guide appropriate treatment, subjective questionnaires, such as the Freezing of Gait Questionnaire (FOGQ) and the New Freezing of Gait Questionnaire (NFOGQ), are commonly used (13; 14). Although these questionnaires may be sufficient to identify the presence of FOG, they are insufficient to objectively describe patients’ FOG severity, and capture treatment effects, as they suffer from recall bias (15), in which the patients may not have been completely aware of their freezing severity, frequency, or impact on daily life. These questionnaires are also poor for intervention studies due to the large test-retest variability resulting in extremely minimal detectable change values. To objectively assess FOG severity, PD patients are asked to perform brief and standardized FOG-provoking tasks in clinical centers. Common tasks include timed-up-and-go (TUG) (16), 180 or 360 degrees turning while walking (17), and 360-degree turning-in-place (360Turn) (18). The TUG is commonly used in clinical practice since the task includes typical everyday motor tasks such as standing, walking, turning, and sitting. In combination with a cognitive dual-task, it has proven to provoke FOG reliably (19). Recently, the 360Turn with a cognitive dual-task was also shown to be practical and reliable to provoke FOG for investigating therapeutic effects on FOG (20). Adding a cognitive dual-task to both the TUG and 360Turn test can increase the cognitive load on individuals, which can result in more FOG events, making these tests more sensitive and perhaps relevant measures of FOG severity in real-life situation (19; 20; 17).

The current gold standard to assess FOG severity during the standardized FOG-provoking tasks is via a post-hoc visual analysis of video footage (21; 17; 22). This protocol requires experts to label FOG episodes and the corresponding FOG manifestations frame by frame (22). Based on the frame-by-frame annotations, semi-objective FOG severity outcomes can be computed, such as the number of FOG episodes (#FOG) and the percentage time spent frozen (%TF), defined as the cumulative duration of all FOG episodes divided by the total duration of the walking task (23). However, this procedure relies on time-consuming and labor-intensive manual annotation by trained clinical experts. Moreover, the inter-rater agreement between experts was not always strong (23), and the annotated #FOG between raters could also contain significant differences due to multiple short FOG episodes being pooled into longer episodes (20).

As a result, there is an interest in automated and objective approaches to assess FOG (5; 24; 25; 26; 27). Apart from traditional automatic approaches that detect freezing segments based on a predefined threshold for high-frequency spectra of the leg acceleration (28), DL approaches were developed to label FOG accurately and objectively using marker-based 3D motion capture (MoCap) data (27). However, marker-based MoCap is cumbersome to set up and is constrained to lab environments. As a result, inertial measurement units (IMU), due to the better portability, were often used to capture motion signals both in a lab and at home (29; 30) and were widely used for automatic FOG assessments (31; 32; 33; 24). The current state-of-the-art DL model, termed multi-stage temporal convolutional neural network (MS-TCN) (34), was initially proposed for frame-by-frame sequence mapping in computer vision tasks. The MS-TCN architecture first generates an initial prediction with several temporal convolution layers and refines the prediction over multiple stages to reduce over-segmentation error (35). Previous studies have shown that MS-TCN exceeded other DL models when dealing with IMU data (35; 36; 37) and shown to be valid for FOG assessment on 3D MoCap data (27). However, MS-TCN is not yet applied to FOG detection with IMU data, which forms the first research gap.

Previous studies proposed automatic FOG detection models for FOG assessment in clinical settings by training and evaluating DL models on datasets that include multiple standardized FOG-provoking tasks measured during both On- and Off-medication states (38; 33; 24; 27). However, seeing the widespread clinical use of the 360Turn task for FOG detection, it is still uninvestigated if DL models can adequately detect FOG in this task, which forms the second gap. Moreover, gait patterns and FOG severity can vary substantially among different FOG-provoking tasks (39) and medication states (40; 41). As a result, whether a DL model can reliably assess FOG in an unseen FOG-provoking task and an unseen medication state is the third research gap.

Additionally, although these standardized FOG-provoking tasks include walking and turning movements, similar to movements in real-life conditions, they do not include sudden volitional stops, which frequently occur during daily activities at home. Hence, it becomes crucial to be able to distinguish between FOG and volitional stops when transitioning toward at-home FOG assessment. These volitional stops usually do not include any lower limb movements and are often considered challenging to distinguish from akinetic freezing (42). Although a previous study proposed using physiological signals to detect discriminative features for classifying FOG from voluntary stops (43), methods using motor signals to distinguish FOG from stops were seldom investigated. To the best of our knowledge, only limited studies proposed FOG detection or prediction on trials with stops using IMU signals (33; 44). However, these studies did not discuss the effect of stopping on FOG detection performance, forming the fourth research gap.

To address the aforementioned gaps, this paper first extends the MS-TCN model to enable automatic FOG assessment on two standardized FOG-provoking tasks (i.e. the TUG task and the 360Turn task) based on IMUs rather than MoCap-based devices. Next, we evaluated whether a DL model trained on TUG tasks could detect FOG in 360Turn tasks and vice versa. Similarly, we investigated whether a DL model trained on Off-medication trials could detect FOG in On-medication trials and vice versa. Finally, we investigated the effect of including or excluding stopping periods on detecting FOG by introducing self-generated and researcher-imposed stopping during standardized FOG-provoking tests. Both self-generated and researcher-imposed stops are hereinafter simply referred to as “stopping”. To this end, the contribution of the present manuscript is four-fold: (1) We extend the state-of-the-art MS-TCN model for fine-grained FOG detection on IMU data. (2) We show, for the first time, that FOG can be automatically assessed during the 360Turn task. (3) We show that DL models cannot generalize to an unseen FOG-provoking task, thereby highlighting the importance of expressive training data in the development of FOG assessment models. (4) We show that the DL model can assess FOG severity with a strong agreement with experts across FOG-provoking tasks and medication states, even in the presence of stopping.

## 1 Methods

### 1.1 Dataset

We recruited 12 PD patients in this study. Subjects were included if they subjectively reported having at least one FOG episode per day with a minimum duration of five seconds. All subjects were asked to fill in the Mini-Mental State Examination (MMSE) (45), Unified Parkinson’s Disease Rating Scale (UPDRS) (46) I to IV, and Hoehn & Yahr (H&Y) Scale (47) for clinical assessments. The inclusion criterion was chosen to maximize the chance of capturing FOG in the lab-based assessment procedure.

All subjects performed a TUG with turning to both directions and a one-minute alternating 360Turn test during the assessments. In the TUG, participants were instructed to stand up from a chair, walk towards a mark placed 2.5 meters from the chair, turn around the mark, walk back to the chair, and sit down. The TUG was performed by all subjects with both left and right turning (two trials). In the 360Turn, participants had to perform rapid alternating 360-degree turns in place for one minute (20). While measuring the standardized FOG-provoking tasks, we included the dual task to provoke more FOG episodes (19; 20). The dual-task consisted of the auditory Stroop task (48; 20), in which the words “high” and “low” were played from a computer with both a high and low pitch voice. Participants were instructed to name the pitch they heard and not repeat the word. As a result, the TUGs and 360Turn tests were grouped into one block (two TUG trials and one 360Turn trial). Each block of tests was measured with and without a dual task (6 trials). We also included measurements containing a self-generated or researcher-imposed stopping period to collect data for further training. Each block with TUG and 360Turn tasks was measured with a stopping period, in which TUGs were performed four times, twice with a stop in the straight walking part and twice with a stop in the turning part of the TUG; while 360Turn was performed one time. The block was repeated with self-generated and researcher-imposed stopping (10 trials). All pre-mentioned assessments were done first in the clinical Off-medication state (at least 12 hours after the last PD medication intake) and repeated in the same order during the On-medication state (at least one hour after medication intake), resulting in 32 trials for each subject. The blocks of all tests were performed in randomized order to counter potential fatiguing effects leading to more FOGs in the last tests.

All participants were equipped with five Shimmer3 IMU sensors attached to the pelvis and both sides of the tibia and talus. All IMUs recorded at a sampling frequency of 64 Hz during the measurements. RGB videos were captured with an Azure Kinect camera at 30 frames per second for offline FOG annotation purposes. For synchronization purposes, triggered signals were sent at regular intervals of one second from the camera to an extra IMU, synced with the other five IMUs. FOG events were visually annotated at a frame-based resolution by a clinical expert, after which all FOG events were verified by another clinical expert using Elan annotation software (22). Annotators used the definition of FOG as a brief episode with the inability to produce effective steps, and the episode ended only when at least two gait cycles were observed (1; 22). Unlike previous studies that considered shuffling as one of the FOG manifestations (1; 5), this study adopts a stricter definition of FOG that distinguishes shuffling and festination as non-FOG events that are related to FOG due to the presence of continuously small steps during walking. During model training and testing, these FOG-related events were considered non-FOG events.

### 1.2 Problem definition

An IMU trial can be represented as 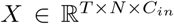, where *T* is the number of samples, *N* specifies the number of IMUs, and *C*_*in*_ is the input feature dimension. Each IMU trial *X* is associated with a ground truth label vector *Y* ∈ ℝ^*T ×L*^, where *L* is the number of output classes, i.e., 2 for non-FOG and FOG. To generate predictions for each sample, the DL model learns a function *f* : *X* → *Y* that transforms a given input sequence *X* = *x*_0_, …, *x*_*T −*1_, where 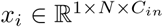, into an output sequence *Ŷ* = *ŷ*_0_, …, *ŷ*_*T−*1_, where *y*_*i*_ *∈ ℝ*^*L*^ that closely resembles the manual annotations *Y*. Generically, the problem formulated thus far is called action segmentation in the computer vision literature (49; 34).

### 1.3 Deep neural network architecture

The state-of-the-art MS-TCN model (34), initially developed for action segmentation based on video data, was extended to IMU sensor data (36; 37) for human activity recognition and 3D MoCap data (27; 35) for FOG assessment. This study, therefore, considered the MS-TCN model as state-of-the-art and applied it to IMU-based FOG assessment.

As shown in figure 1, there are two significant blocks in our MS-TCN model: (1) an initial prediction block with a single-stage temporal convolutional network (TCN) to generate initial annotations from the input IMU signals; and (2) a prediction refinement block with multiple stages of TCN for incremental refinements that reduces the number of segmentation error (34; 35). The characteristics of the two blocks are further discussed in this section.

**Figure 1:**
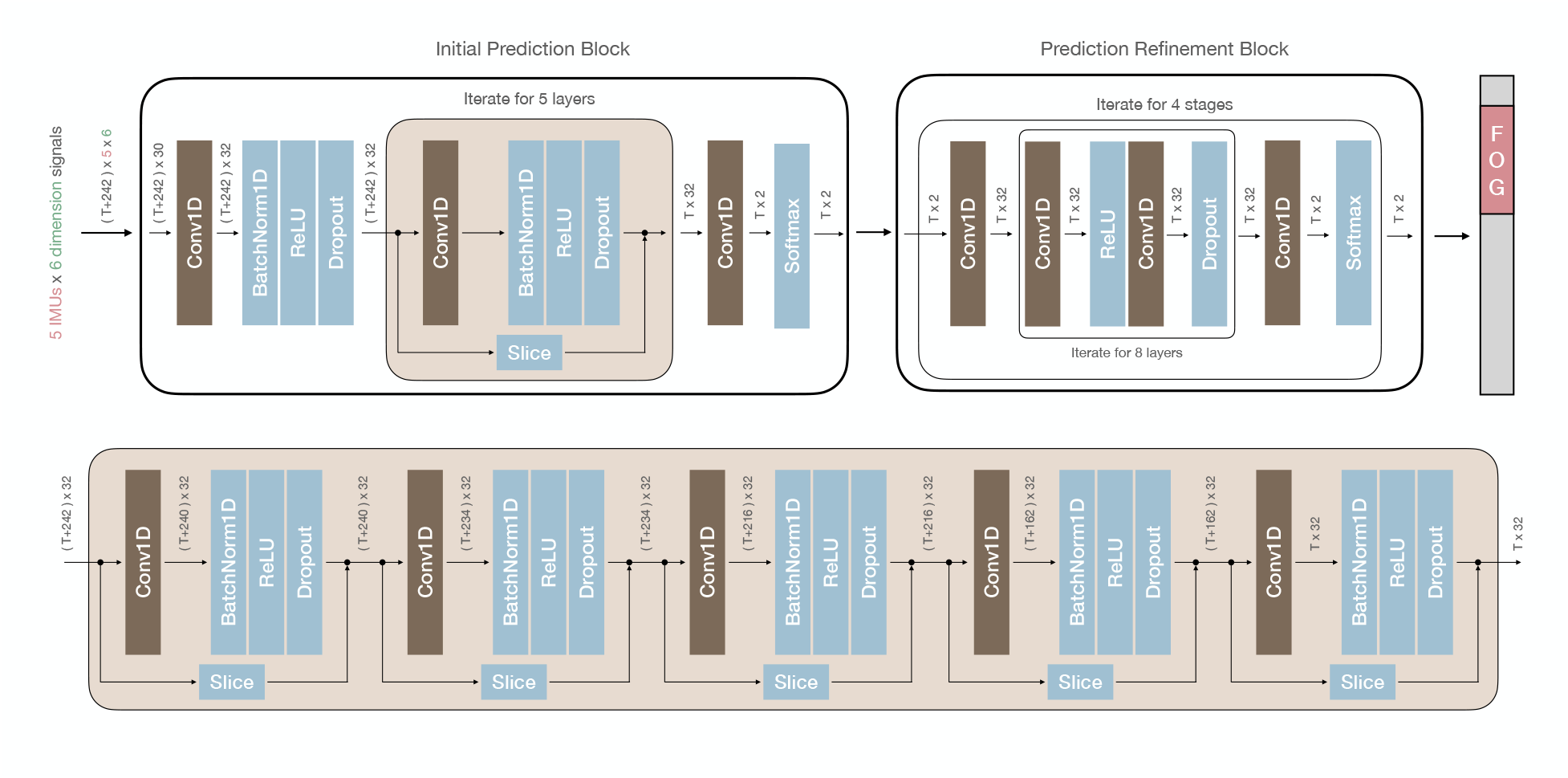
Overview of the MS-TCN architecture and the single-stage TCN with a many-to-one architecture in the initial prediction block

Our modified MS-TCN generates an initial prediction with a single-stage TCN with five temporal convolution layers during training time and refines the predictions over four stages of TCN, each with eight temporal convolution layers. The input consists of six-dimensional signals with *T* samples for five IMUs, and it was zero padded with 121 frames both in the front and end. The model predicts the probability of positive (freezing) and negative (non-freezing) for each frame of the input sequence. The number of hidden features was set to 32. The temporal convolution layers in the initial prediction block increase the receptive field exponentially by a factor of 3, resulting in a receptive field of 243 frames. Due to the many-to-one prediction, valid convolutions with a kernel size of 3 decrease the output samples of each layer from *T* + 242 to *T* + 240 and so on to *T*. The temporal convolution layers in the refinement block applied the same convolution, in which the input and output sequence contained the same number of samples, with the input sequence of each temporal convolution layer zero-padded on both sides.

#### 1.3.1 Initial prediction block

TCN was initially designed to model long-range patterns in a time sequence (49). The network can take a sequence of any length and map it into an output sequence of the same length, often described as a many-to-many scheme. With multiple layers of convolutional filters, a TCN extracts relevant features from the input data (49). The filters operate on small windows of the input data, called the receptive field, and produce outputs combined and passed through non-linear activation functions to form a final output. The kernel size of the convolution filter determines the number of time steps each filter will consider when computing its output, which also determines the receptive field. In regular convolutions, the filter is typically moved one sample at a time. In strided convolutions, a stride parameter determines the step size with which the filter moves across the input data, reducing the spatial or temporal output representation for computational efficiency. To increase the receptive field of the model with fewer parameters, dilated convolutions were widely used by adding gaps, determined by the dilation factor, between the elements of the convolutional filters (50; 34). Based on the number of output samples, convolutions are divided into two types: valid and same. Valid convolution involves sliding the filter over each unique position within the input without padding, resulting in a reduction of the temporal representation. On the other hand, the same convolution involves padding with zeros so that the size of the output representation remains the same as the input.

The initial prediction block in the MS-TCN model contains a single-stage TCN to generate initial frame-by-frame FOG detections given an IMU trial. Given a sequence of *T* samples: *X* = *x*_0_, *x*_1_, …, *x*_*T −*1_, the model will predict an output sequence of *T* samples: *Ŷ* = *ŷ*_0_, *ŷ*_1_, …, *ŷ*_*T −*1_. With the many-to-many scheme, the model is trained on sequences with a receptive field of *n* samples and learns to generate an output sequence of *n* samples. However, previous studies have shown that the many-to-one training strategy (51), in which the DL model is trained on input sequences of length *n* with an output prediction for only a single sample, results in better generalization (52; 51). As a result, we introduced the many-to-one training scheme to the initial TCN block of the MS-TCN model. With the many-to-one architecture, during training, all input sequences were divided into multiple chunks with a receptive field of size *n*, each with a single-sample prediction, with the annotation of the middle sample in each chunk as the ground truth. During inference, the entire input sequence was processed for faster generation by sharing the intermediate states of each node in the neural network. Specifically, given an input sequence with length *T* and a receptive field of size *n*, the input sequence length will reduce from *T* to *T −* (*n −* 1) after prediction. To avoid losing frames, a padding scheme was applied by replicating the boundaries of the input sequence with the corresponding receptive field size, resulting from the original input of length *T* to *T* + (*n −* 1). For example, as shown in figure 2, when the receptive field was set to 27 for an input trial of length *T*, the whole sequence was padded with size 13 on both sides. After prediction, the model will produce a prediction for each sample, with a size of *T*. This study padded all input sequences with zero values on both sides, given the corresponding receptive field size.

**Figure 2:**
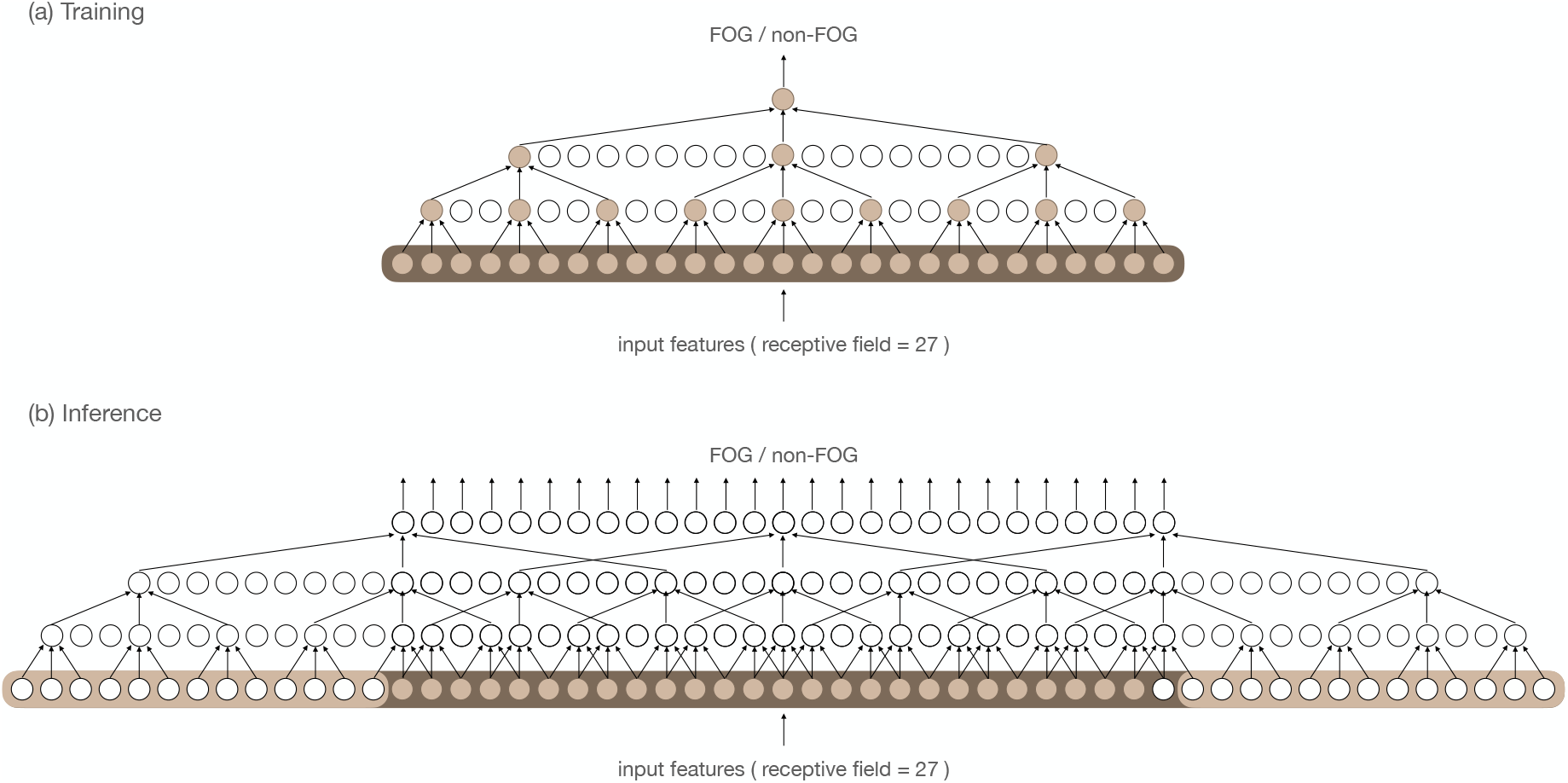
Comparison between the TCN in the initial prediction block during training and inference

This toy example is shown for a receptive field of 27 frames. The many-to-one training scheme was applied for better generalization and to increase memory usage (52; 51). During training, a sequence of inputs equal to the receptive field (here 27) was mapped into a single output sample. During inference, the variable length sequence, equal to the number of samples within a FOG-provoking task (here 27), was zero padded by half the receptive field minus 1 (here 13) to generate predictions for each sample.

#### 1.3.2 Prediction refinement block

After generating initial predictions from the initial TCN block, the prediction refinement block with multiple TCN stages was introduced to reduce over-segmentation errors in the initial predictions, which occur when generating predictions on a frame-by-frame basis (34; 35) and negatively impact FOG outcomes (27). The prediction refinement block of the MS-TCN architecture (34) was designed to perform a many-to-many sequence prediction. Given a sequence of *T* samples generated from the initial prediction block, each stage of the refinement block will refine the prediction of the previous stage by generating an output sequence of *T* frames. All the predictions generated from each stage are then used to calculate the loss function during training time. Figure 1 illustrates the changes in the sequence length from the initial prediction block to the refinement block and the final prediction.

#### 1.3.3 Loss functions

This paper applied the same loss function as in (34), which utilized a combination of a classification loss and an additional smoothing loss to reduce over-segmentation errors during training. The cross-entropy (CE) loss was selected as the classification loss, defined as:

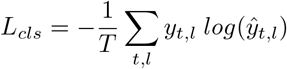

where *T* is the number of samples and *L*_*cls*_ is the CE loss with *y*_*t,l*_ and *ŷ*_*t,l*_ the ground truth label and predicted probability for the class *l* at time *t*, respectively.

FOG generally occurs less frequently than non-FOG during in-lab measurements (20), which lead to class-imbalance problem when training DL (53). To tackle the class-imbalance problem between the freezing and non-freezing class ratio, a weighted factor *α* was introduced to the loss function as a hyperparemeter (54), resulting in the weighted CE loss, defined as:

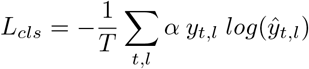

To train the initial prediction block, due to the many-to-one training strategy, only the classification loss, CE loss (*L*_*cls*_), of each mini-batch is minimized. For the multiple refinement stages, with a total number of *S* stages, the sum of the losses (*L*) over all stages *s* is minimized, defined as:

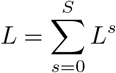

where *L*^*s*^ is the loss stage *s*. To reduce over-segmentation error, *L*^*s*^ is a combination of the CE loss and a smoothing loss, with a hyperparameter *λ* that controls the contribution of each loss function, defined as:

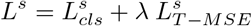

The smoothing loss *L*_*T −MSE*_ is a truncated mean squared error of the sample-wise log-probabilities:

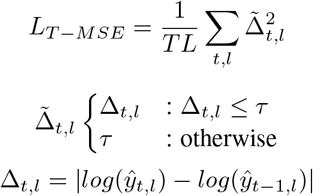

where *T* is the sequence length, *L* is the total number of classes, and *ŷ*_*t,l*_ is the probability of class *l* at sample *t*, and *τ* is the hyperparameter that defines the threshold to truncate the smoothing loss. The larger the *τ*, the smoothing loss penalizes more of the discontinuity, which reduces the model’s capability in detecting the true boundaries between different classes (34).

#### 1.3.4 Implementation details

The hyperparameters were selected based on previous studies that proposed the DL model with the many-to-one architecture (51) and the MS-TCN model (34). As shown in figure 1, the initial prediction block follows the design architecture of (51). Five convolutional layers were included in the single-stage TCN; each increases the receptive field exponentially by a factor of three, resulting in a receptive field with 243 samples (around 4 seconds). The refinement block follows the design architecture of (34). The number of refinement stages was set to four (*S* = 4), each stage is a TCN with the number of layers within set to eight, and the dilation factor doubled at each layer, i.e., 1, 2, 4, …, 128, with a kernel size of three, the receptive field of the refinement block is 511 samples (around 8 seconds).

We applied the same hyperparameters for the initial TCN and refinement blocks to maintain consistent settings. The number of filters was set to 32, and the kernel size of all dilated temporal convolutions was equal to 3. ReLU activations were included after each block. For the CE loss, the weighted term *α* of each class was set to be the inverse of the frequency of each class. For the smoothing loss, *τ* was set to 4, and *λ* was set to 0.15. All experiments used the Adam optimizer (55) with a learning rate of 0.0005 and decay with a factor of 0.95 for each epoch. The beta1 and beta2 parameters in Adam were set to 0.9 and 0.999. All models were trained for 50 epochs, as all models converged over this value. The initial TCN block was trained with a batch size of 1024 (1024 chunks under the many-to-one scheme), while the refinement block was trained with a batch size of one (one IMU trial).

Before training and testing the model, all six channels of the IMU signals for each trial were centralized by subtracting the mean value of each signal to remove the constant bias.

### 1.4 Evaluation

To evaluate the performance of the model, datasets were partitioned using a leave-one-subject-out (LOSO) cross-validation approach. The LOSO cross-validation approach iteratively splits the data according to the number of subjects in the dataset. One subject is evaluated, while the others are used to train the model. This procedure was repeated until all subjects had been used for evaluation. This approach mirrored the clinically relevant scenario of FOG assessment in newly recruited subjects (56), where the model assesses FOG in unseen subjects. The result for all models shown in this study were averaged over all unseen subjects using the LOSO cross-validation approach.

#### 1.4.1 Experimental settings

##### Clinical setting

To support FOG assessment in clinical settings, which typically do not include stopping, this study first investigated the overall and relative performance of a generic model trained across standardized FOG-provoking tasks that do not include stopping. Next, we assessed generalization across FOG-provoking tasks and medication states by studying the effect of including or excluding training data from a specific task or medication state on detecting FOG.

##### Towards the home setting

To move towards FOG assessment in daily life where stopping frequently occurs, we trained and evaluated the performance of a generic model trained across trials with stopping. Next, we assessed the effect of including or excluding stopping periods on detecting FOG.

##### Naming convention

The naming convention of all the DL models that were evaluated in this study with their corresponding training data is shown in table 1. The generic model trained for clinical measurements (i.e., excluding stopping) was termed “Model_Clinical”. Models trained with less data variety were termed (i.e., trained for a specific task or medication state): “Model_TUG”, “Model_360Turn”, “Model_Off”, and “Model_On”. The generic model trained to work towards FOG assessment in daily life (i.e., including stopping) was termed “Model_Stop”. To compare the effect of stopping, we evaluated Model_Clinical and Model_Stop. In order to maintain a similar amount of FOG duration in the trained data, Model_Stop was trained on trials that included stopping but not those that excluded it.

**Table 1:** Naming convention of the deep learning models evaluated in this study with their corresponding training data

| Usage | Model Name | FOG-provoking task |  | Medication state |  | Stopping |  |
| --- | --- | --- | --- | --- | --- | --- | --- |
|  |  | TUG | 360Turn | Off | On | Exclude | Include |
| FOG detection in clinical practice | Model_TUG | v |  | v | v | v |  |
|  | Model_360Turn |  | v | v | v | v |  |
|  | Model_Off | v | v | v |  | v |  |
|  | Model_On | v | v |  | v | v |  |
|  | Model_Clinical | v | v | v | v | v |  |
| Towards FOG detection in daily life | Model_Stop | v | v | v | v |  | v |

This study extended the state-of-the-art MS-TCN model for IMU-based FOG detection. Models trained for specific tasks or medication states for standardized measurements were termed “Model\_TUG”, “Model\_360Turn”, “Model\_Off”, and “Model\_On”. A DL model trained for FOG assessment in the clinical centers, trained on standardized tasks excluding stops, was termed “Model\_Clinical”. A DL model trained to work towards FOG assessment in daily life, trained on standardized tasks including stops, was termed “Model\_Stop”.

#### 1.4.2 Metrics

From a clinical perspective, FOG severity is typically assessed in terms of percentage time-frozen (%TF), and the number of detected FOG episodes (#FOG) (23). This paper used %TF as the primary outcome and #FOG as a secondary outcome based on previous studies (20; 24). To assess the agreement between the model predictions and the expert annotations for each of the two clinical metrics, we calculated the intra-class correlation coefficient (ICC) with a two-way random effects analysis (random trials, random raters) (ICC(2,1)), in which both the raters and the subjects are treated as random effects, meaning that they are assumed to be a random sample from a larger population (57). The ICCs between the model and experts were calculated subject-based, with one %TF and #FOG per subject. In other words, the %TF and #FOG were calculated over all trials for each subject. The strength of the agreement was classified according to (58): ≥0.80: strong, 0.6-0.79: moderately strong, 0.3-0.59: fair, and *<*0.3: poor.

From a technical perspective, the sample-wise F1 score (Sample-F1) is a metric commonly used in classification problems to evaluate the quality of a model’s predictions at the individual sample level. It provides a balanced measure of a model’s ability to identify positive and negative classes. In binary classification, Sample-F1 is computed by comparing the predicted and true labels. Each sample is classified as true positive (TP), false positive (FP), or false negative (FN) by a sample-wise comparison between the experts’ annotation and model predictions. Sample-F1 is calculated under the formula:

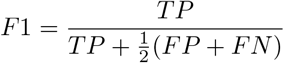

Additionally, the segment-wise F1-score at *k* (Segment-F1@*k*) proposed by (49) is a metric that penalizes over and under-segmentation errors. It allows only minor temporal shifts for the predicted segment, resulting in a much stricter evaluation metric than sample-wise metrics such as Sample-F1 (27). To compute Segment-F1@*k*, action segments are classified as TP, FP, or FN by comparing the intersection over union (IoU) to a pre-defined threshold *k*. The IoU is calculated as the intersection length of the predicted segment and the ground-truth segment divided by the union between the two segments. If the corresponding IoU of a predicted segment is large than *k*, the predicted segment is TP; otherwise, it is FP. All unpaired ground-truth segments are considered FN. Based on previous studies (27; 35), we set the threshold *k* for IoU as 50% (Segment-F1@50). Additionally, an example to compare %TF, #FOG, and Segment-F1@50 is shown in figure 3. The %TF and #FOG for both annotations are 40% and 2 for trial 1, 10% and 1 for trial 2, resulting in a high ICC value of 1. However, the Segment-F1@50 is 0.67 for trial 1 and 0 for trial 2, resulting in an averaged Segment-F1@50 of 0.335. This example shows that although ICC is widely used in previous studies when comparing the inter-rater agreement of %TF and #FOG, it contains the disadvantages of not penalizing shifted annotations, a problem that Segment-F1@50 overcomes. This study calculated one Sample-F1 and Segment-F1@50 for each subject by taking the averaged Sample-F1 and averaged Segment-F1@50 over all trials of that subject. The overall Sample-F1 and Segment-F1@50 under the LOSO cross-validation approach were calculated by averaging the metrics over all subjects.

**Figure 3:**
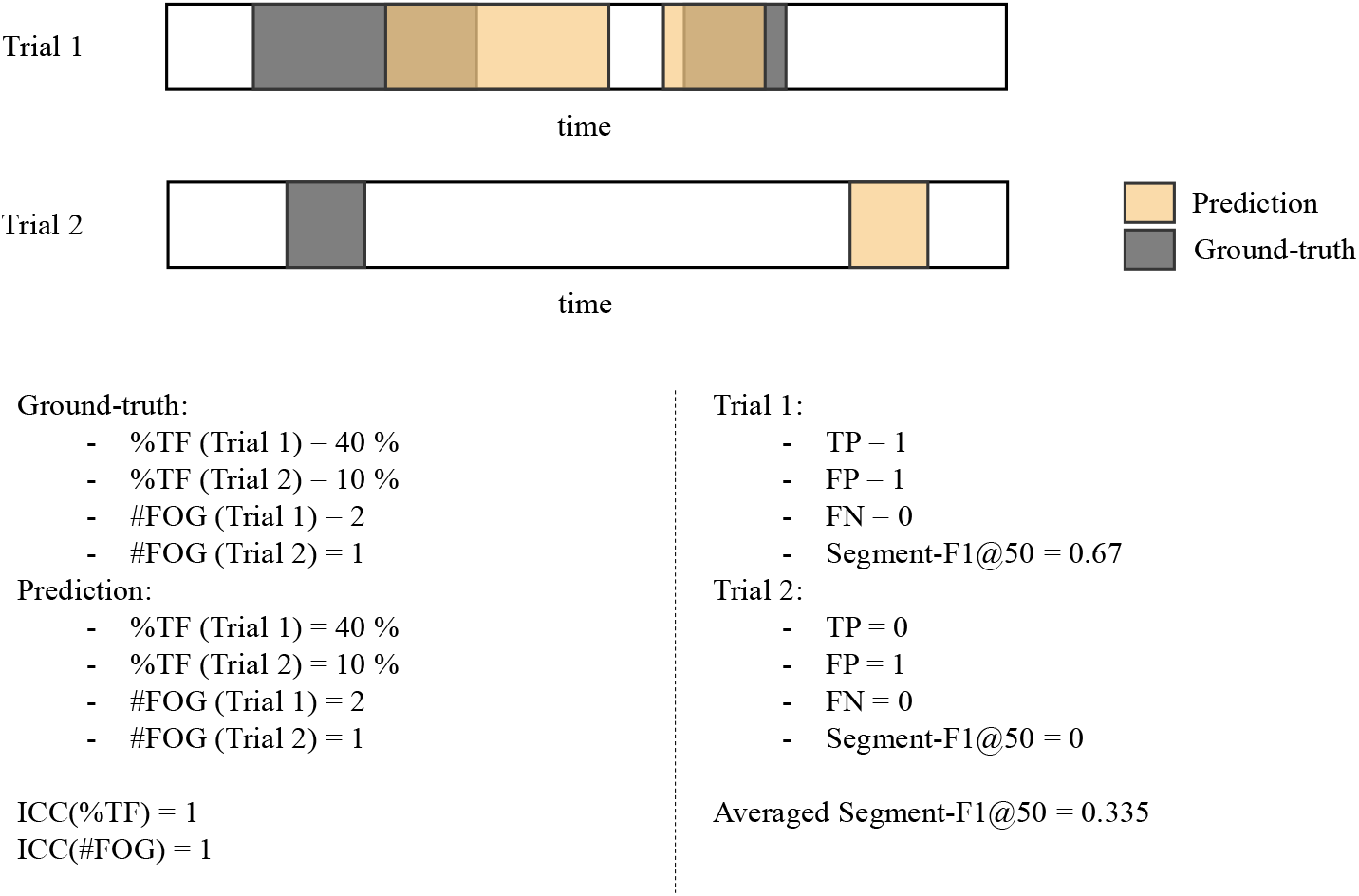
An example for comparing ICC and segment-wise F1 score

Based on the above discussion, when comparing the performance between different models, i.e., Model_TUG vs. Model_360Turn, Model_Off vs. Model_On, and Model_Clinical and _Model_Stop, only Sample-F1 and Segment-F1@50 were used. Whereas when showing the agreement between the two generic models and the experts in terms of FOG severity outcomes, the ICC values for %TF and #FOG were reported.

This toy example shows the annotations on two trials with the ground-truth annotation as gray and the predicted annotation as yellow. The x-axis represents the timeline for the annotations. When calculating the agreement between the ground-truth and prediction, the %TF and #FOG are both 40% and 2 for the first trial and 10% and 1 for the second trial, resulting in an ICC value of 1. On the other hand, for the segment-wise F1@50 of the first trial, since FP=1 (the first FOG segment has an IoU less than 50%), TP=1 (the second FOG segment has an IoU over 50%), and FN=0, resulting in a segment-F1@50 with 0.67. For the second trial, FP=1, TP=0, and FN=0 resulted in a segment-F1@50 with 0. Thus, the mean Segment-F1@50 equals 0.335. This example shows the disadvantage of using the ICC value of %TF and #FOG to measure the alignment between two annotations.

#### 1.4.3 Statistical analysis

The Bland–Altman plot (59) was applied to investigate the systematic bias of the %TF and #FOG between the prediction of Model_Clinical and the experts’ annotation. To investigate whether the difference in Sample-F1 and Segment-F1@50 for each subject between two DL models, i.e., Model_TUG vs. Model_360Turn, Model_On vs. Model_Off, and Model_Clinical vs. Model_Stop, was statistically significant, the paired Student’s t-test (60) was applied, with the number of pairs equal to the number of subjects evaluated with LOSO. The homogeneity of variances was verified in all metrics across subjects with Levene’s tests (61). The Shapiro-Wilk test (62) was used to determine whether the variables were normally distributed across subjects. The significance level for all tests was set at 0.05. All analyses were performed using SciPy 1.7.11, bioinfokit 2.1.0, statsmodels 0.13.2, and pingouin 0.3.12, written in Python version 3.7.11.

## 2 Results

This section first describes the dataset characteristics. Next, we discuss the result of automatic FOG assessments at two levels: (1) FOG detection for clinical measurements with a discussion on the effect of FOG-provoking tasks and medication states, and (2) FOG detection for moving towards daily life with a discussion on the effect of stopping.

### 2.1 Dataset characteristics

Table 2 shows the clinical characteristics of the twelve PD patients. Participants varied in their age (mean: 69.33 years, range: 57*−*76), disease duration (mean: 12.33 years, range: 3*−*23), and self-reported FOG severity with NFOGQ (mean: 20.54, range: 12*−*26).

**Table 2:** Subject characteristics

| | Average ± SD |
| --- | --- |
| Age | 69.33 ± 6.28 |
| PD duration | 12.33 ± 6.25 |
| MMSE (45) | 25.45 ± 8.57 |
| UPDRS I + II + III + IV (46) | 88.16 ± 22.55 |
| H&Y (47) | 2.60 ± 0.80 |

The subject characteristics of all 12 PD patients at the baseline assessment. All characteristics were measured during the On-phase and Off-phase of the medication cycle.

According to table 3, a total of 346 trials were collected. Freezing occurred in 38.43% of trials (133 out of 346 trials), with %TF of 14.62% and #FOG of 530 observed. Based on the dataset measurement protocol, 32 trials were collected for each subject. The number of trials more than 32 was due to repeated measurements, and the number of trials less than 32 was due to technical difficulties.

**Table 3:**
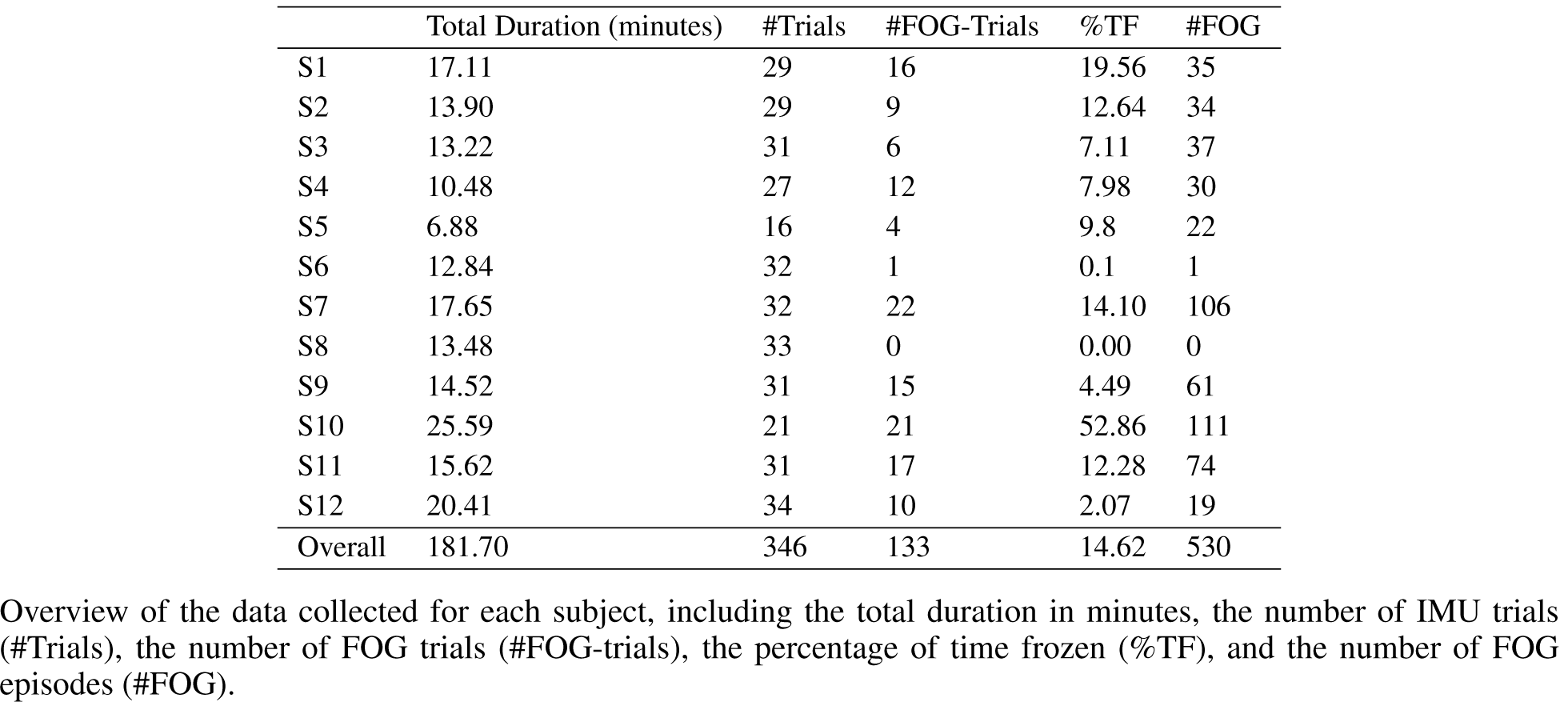
Dataset characteristics

The 346 trials in the dataset included 133 trials (81.11 minutes) collected within the clinical setting, i.e., trials without stopping, and 213 trials (100.60 minutes) with stopping included. According to table 4, all 133 trials without stopping were used to train Model_Clinical, while all 213 trials with stopping were used to train Model_Stop. Within the 133 trials without stopping, 89 TUG trials (35.99 minutes) were used to train Model_TUG, and 44 360Turn trials (45.11 minutes) were used to train Model_360Turn. Similarly, 67 Off-medication trials (45.75 minutes) were used to train Model_Off, and 66 On-medication trials (35.36 minutes) were used to train Model_On. These models were evaluated and discussed in the following sections.

**Table 4:** Overview of the number of trials, total duration (minutes), and FOG outcome of the training data for all models evaluated in this study

| Model Name | #Trials | Total Duration (minutes) | %TF | #FOG |
| --- | --- | --- | --- | --- |
| Model_TUG | 89 | 35.99 | 19.88 | 103 |
| Model_360Turn | 44 | 45.11 | 21.71 | 179 |
| Model_Off | 67 | 45.75 | 27.37 | 205 |
| Model_On | 66 | 35.36 | 12.53 | 77 |
| Model_Clinical | 133 | 81.11 | 20.90 | 282 |
| Model_Stop | 213 | 100.60 | 9.57 | 248 |

A total of 133 trials were measured with standardized FOG-provoking tests excluding stops, which were used to train Model\_Clinical, and 213 trials were measured with self-generated and researcher-imposed stops, which were used to train Model\_Stop. Within the 133 trials, 89 trials of TUG tasks were used to train Model\_TUG, and 44 were used to train Model\_360Turn. Similarly, 67 trials Off medication were used to train Model\_Off, and 66 trials were used to train Model\_On. Trial durations are shown in minutes. FOG severity is quantified by means of the %TF and #FOG.

### 2.2 Clinical setting: FOG detection

This study first trained and evaluated the proposed model trained for FOG detection in standardized clinical setting (i.e., trials without stopping). Model_Clinical detected #FOG per subject varied from 3 to 80, amounting to 335 FOG episodes, while the %TF varied from 0.52% to 70.49%. When comparing with experts’ annotations, the model had a strong agreement in terms of %TF, (ICC=0.92, CI=[0.68,0.98]), and #FOG (ICC=0.95, CI=[0.72,0.99]). The Bland–Altman plots shown in figure 4 revealed a systematic error across FOG severity from the model, with a mean bias of *−*4.06 (CI=[*−*7.41,*−*0.72]) for %TF and *−*4.41 (CI=[*−*7.66,*−*1.17]) for #FOG. For %TF, the limits of agreement (LOA) fall within the range of *−*14.40% (CI=*−*20.19,*−*8.59) to 6.26% (CI=[*−*0.45,12.05]), showing that it was confident that the differences between the model and the experts would lie in the range of *−*14.40% to 6.26%. For #FOG, the LOA fall within the range of *−*14.43 (CI=[*−*20.04,*−*8.80]) to 5.59 (CI=[*−*0.02,11.21]), showing that the differences between the model and the experts will lie in the range of *−*14.43 to 5.59.

**Figure 4:**
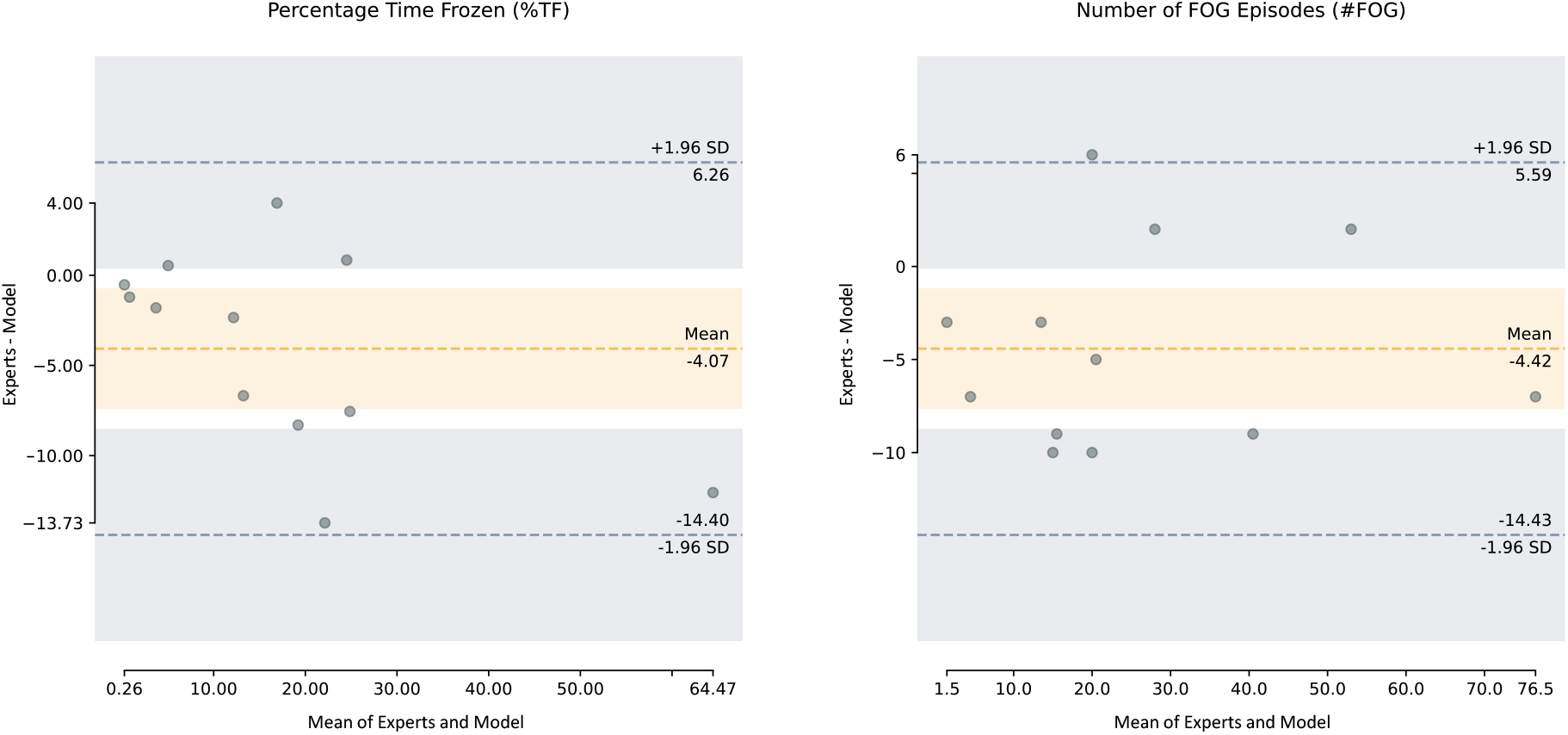
Bland–Altman plot for the clinical metrics from Model_Clinical and the experts

The dots represent the difference in scores per patient on the y-axis (i.e., model’s %TF or #FOG subtracted from experts’ %TF or #FOG), plotted against the mean score per patient from the model and the experts on the x-axis. The orange shaded area represents the 95% CI for the mean bias, and the gray shaded area represents the 95% CI for the upper and lower limits of agreement. A negative mean error indicates that the model overestimates with %TF and #FOG compared with the experts’ annotation.

Additionally, when evaluating all standardized trials (i.e., without stopping) within the dataset, results showed that 56.70% of the false positive (FP) samples were annotated as FOG-related segments, i.e., shuffling and festination, meaning that the model tended to annotate FOG-related samples as FOG. According to the qualitative example of the model and experts’ annotations in figure 5, the model generally predicted broader FOG segments compared to the experts’ annotations, resulting in an overestimation of %TF. Also, the model tends to split some experts’ annotated FOG segments into two different FOG segments, resulting in overestimating #FOG.

**Figure 5:**
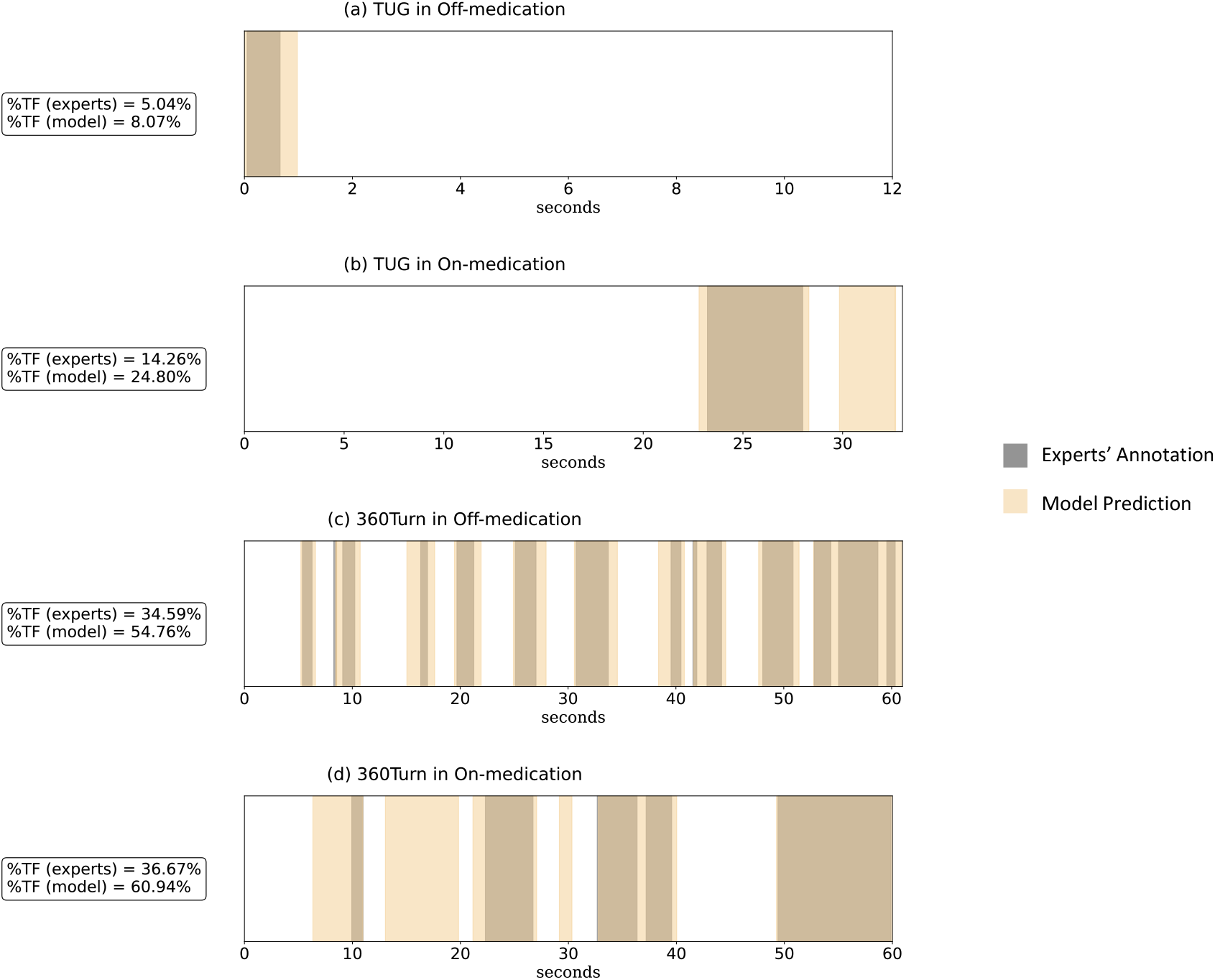
Overview of the annotations for four typical IMU trials from two patients

Four typical trials include annotations for IMU trials measured during four settings: a) TUG in Off-medication (S3), b) TUG in On-medication (S1), c) 360Turn in Off-medication (S3), d) 360Turn in On-medication (S1). The figures visualize the difference between the manual FOG segmentation by the clinician and the automated FOG segmentation by the MS-TCN model. The x-axis denotes the time of the trial in seconds. The gray region indicates the experts’ annotated FOG, and the yellow region indicates the model-annotated FOG. The color gradient visualizes the overlap or discrepancy between the model and experts’ annotations. The figure shows that the model generally annotated broader FOG events compared to experts’ annotation, resulting in a systematic error in %TF shown in figure 4.

Next, we assessed the relative performance of the generic model in detecting FOG for trials with a specific FOG-provoking task, medication state, or with and without stopping. As shown in table 5, Model_Clinical had a strong agreement with the experts in terms of %TF (all ICCs>0.92) and #FOG (all ICCs>0.84). Results showed that it was more difficult for the model to detect FOG in 360Turn tests than TUG in terms of the average Segment-F1@50 (360Turn: 0.45; TUG: 0.67) and Sample-F1 (360Turn: 0.58; TUG: 0.72). Similarly, it was more difficult for the model to detect FOG in Off trials than On trials (Segment-F1@50: 0.55 vs. 0.64; Sample-F1: 0.65 vs. 0.69). However, these results were not reflected in the ICC values for %TF and #FOG, which also shows the inadequate ability of such metrics when comparing different models.

**Table 5:** Overview of the performance of Model_Clinical and Model_Stop

| Model | Test data | ICC(%TF) | ICC(#FOG) | Segment-F1@50 | Sample-F1 |
| --- | --- | --- | --- | --- | --- |
| Model Clinical | TUG | 0.95, CI=[0.86,0.99] | 0.97, CI=[0.89,0.99] | 0.67 | 0.72 |
|  | 360Turn | 0.94, CI=[0.78,0.98] | 0.84, CI=[0.57,0.95] | 0.45 | 0.58 |
|  | Off-state | 0.94, CI=[0.74,0.99] | 0.94, CI=[0.76,0.99] | 0.55 | 0.65 |
|  | On-state | 0.92, CI=[0.76,0.98] | 0.90, CI=[0.70,0.97] | 0.64 | 0.69 |
|  | All trials | 0.92, CI=[0.68,0.98] | 0.95, CI=[0.72,0.99] | 0.60 | 0.67 |
| Model Stop | TUG | 0.78, CI=[0.42,0.93] | 0.77, CI=[0.41,0.93] | 0.65 | 0.67 |
|  | 360Turn | 0.98, CI=[0.90,1.00] | 0.65, CI=[0.16,0.89] | 0.33 | 0.47 |
|  | Off-state | 0.89, CI=[0.52,0.97] | 0.83, CI=[0.53,0.95] | 0.55 | 0.62 |
|  | On-state | 0.98, CI=[0.96,1.00] | 0.69, CI=[0.22,0.91] | 0.62 | 0.64 |
|  | All trials | 0.95, CI=[0.73,0.99] | 0.79, CI=[0.46,0.94] | 0.59 | 0.63 |

We proceeded to evaluate how Model_Clinical performed in comparison to models specifically designed for various FOG-provoking tasks and medication conditions, including Model_TUG, Model_360Turn, Model_Off, and Model_On. When testing on TUG trials, there was no difference between Model_Clinical and Model_TUG in terms of Segment-F1@50 (p=0.726) and Sample-F1 (p=0.641). When testing on 360Turn trials, there was no difference between Model_Clinical and Model_360Turn in terms of Segment-F1@50 (p=0.149) and Sample-F1 (p=0.331). Similarly, When testing on Off-medication trials, there was no difference between Model_Clinical and Model_Off in terms of Segment-F1@50 (p=0.545) and Sample-F1 (p=0.892). When testing on On-medication trials, there was no difference between Model_Clinical and Model_On in terms of Segment-F1@50 (p=0.483) and Sample-F1 (p=0.184). These results suggest that Model_Clinical can generalize well across different FOG-provoking tasks and medication states for FOG detection in clinical measurements, and there is no significant difference between the model and the models that were trained for specific conditions.

#### 2.2.1 Effect of FOG-provoking tasks and medication states

Next, we investigated the effect of including or excluding data from specific tasks or medication states. As shown in table 6, when testing on TUG trials, Model_TUG resulted in a statistically higher Segment-F1@50 (p<0.005) and Sample-F1 (p<0.005) than Model_360Turn. Similarly, when testing on 360Turn trials, Model_360Turn resulted in a higher Segment-F1@50 and Sample-F1 than Model_TUG, though the differences were not statistically significant. On the other hand, when testing on Off-medication trials, no difference was found between Model_Off and Model_On in terms of Segment-F1@50 (p=0.952) and Sample-F1 (p=0.957). Similarly, when testing on On-medication trials, no difference was found between Model_Off and Model_On in terms of Segment-F1@50 (p=0.579) and Sample-F1 (p=0.307). The results showed that DL models trained only on TUG trials could still detect FOG in 360Turn trials, while DL models trained only on 360Turn could not detect FOG in TUG trials. In contrast, DL models trained without trials for specific medication states could detect FOG on trials measured during unseen medication states. In other words, the data variance between different FOG-provoking tasks was more challenging to model than between different medication states.

**Table 6:** Model comparison results in terms of training on different FOG-provoking task or medication state

| Model | Test data | Segment-F1@50 |  | Sample-F1 |  |
| --- | --- | --- | --- | --- | --- |
|  |  | Mean | t-test (#pair) | Mean | t-test (#pair) |
| Model_TUG | TUG | 0.70 | t(12)=6.19,p<0.005 | 0.75 | t(12)=6.07,p<0.005 |
| Model_360Turn | TUG | 0.10 |  | 0.15 |  |
| Model_360Turn | 360Turn | 0.53 | t(12)=2.14,p=0.055 | 0.62 | t(12)=1.77,p=0.104 |
| Model_TUG | 360Turn | 0.44 |  | 0.56 |  |
| Model_Off | Off | 0.52 | t(12)=-0.06,p=0.952 | 0.66 | t(12)=0.05,p=0.957 |
| Model_On | Off | 0.53 |  | 0.65 |  |
| Model_On | On | 0.68 | t(11)=0.57,p=0.579 | 0.76 | t(11)=1.07,p=0.307 |
| Model_Off | On | 0.65 |  | 0.71 |  |

We investigated the generalization of task-specific models to unseen tasks and the generalization of medication-specific models to unseen medication states. The third and fourth column depicts Segment-F1@50 averaged over all subjects and the paired t-test result. The fifth and sixth columns depict the Sample-F1 averaged over all subjects and the paired t-test result. The number of subjects (pairs) was 12 for TUG, 360Turn, and Off-state, while only 11 subjects were considered for On-state due to technical problems for subject 5 during On-medication state measurements.

### 2.3 Towards the home setting: FOG detection

To move towards FOG detection in daily life, we trained and evaluated the DL model, Model_Stop, on trials collected with stopping. As shown in table 5, when comparing with experts’ annotations, Model_Stop had a strong agreement in terms of %TF, (ICC=0.95, CI=[0.73,0.99]), and a moderately stong agreement in terms of #FOG (ICC=0.79, CI=[0.46,0.94]). Similar to FOG detection in clinical settings, results show that it was also more difficult for the model to detect FOG in 360Turn tests than TUG in terms of the average Segment-F1@50 (360Turn: 0.33; TUG: 0.65) and Sample-F1 (360Turn: 0.47; TUG: 0.67). Also, it was more difficult for the model to detect FOG in Off trials than On trials (Segment-F1@50: 0.55 vs. 0.62; Sample-F1: 0.62 vs. 0.64).

#### 2.3.1 Effect of stopping periods

Next, we investigated the effect of stopping periods on FOG detection by comparing the performance of DL models trained on trials with and without self-generated and researcher-imposed stopping, i.e., Model_Clinical and Model_Stop. According to the results shown in table 7, when evaluating trials collected during standardized measurements, i.e., trials without stopping, there was no difference found between Model_Clinical and Model_Stop in terms of Segment-F1@50 (p=0.550) and Sample-F1 (p=0.326). The results revealed that adding a stopping period in trials during training does not affect the performance of the model in detecting FOG, as both p-values are larger than 0.05, meaning there was no difference between Model_Clinical and Model_Stop.

**Table 7:** Model comparison results in terms of training with/without trials containing stops

| Model | Test data | Segment-F1@50 |  | Sample-F1 |  | #FP-Stop |
| --- | --- | --- | --- | --- | --- | --- |
|  |  | Mean | t-test (#pair) | Mean | t-test (#pair) |  |
| Model_Clinical | Non-stop | 0.60 | t(12)=0.16,p=0.874 | 0.67 | t(12)=0.02,p=0.979 |  |
| Model_Stop | Non-stop | 0.59 |  | 0.67 |  |  |
| Model_Clinical | Stopping | 0.40 | t(12)=-5.31,p<0.005 | 0.46 | t(12)=-4.39,p<0.005 | 74/210 |
| Model_Stop | Stopping | 0.59 |  | 0.63 |  | 16/210 |

We investigated the effect of including or excluding stopping periods in FOG detection by comparing models trained with (i.e., Model\_Stop) and without stopping trials (i.e., Model\_Clinical). The third and fourth column depicts Segment-F1@50 averaged over all 12 subjects and the paired t-test result. The fifth and sixth columns depict the Sample-F1 averaged over all 12 subjects and the paired t-test result. The seventh column depicts the number of stops detected as FOG with respect to the total number of stops (#FP-Stop). A black box was added for #FP-Stop when testing on trials without stopping as it would not be possible to detect stopping as FOG.

When evaluating trials collected with stopping periods, the Segment-F1@50 for Model_Stop (mean=0.60) was significantly higher than Model_Clinical (mean=0.39; p=0.005). Similarly, the Sample-F1@50 for Model_Stop (mean=0.65) was significantly higher than Model_Clinical (mean=0.44; p<0.005). Additionally, among the 210 observed stops within the dataset, only 16 were mislabeled as FOG from Model_Stop, while 74 were annotated as FOG from Model_Clinical. The results indicated that the model trained with trials that include stopping could learn to differentiate stopping from FOG, resulting in a statistically higher Segment-F1@50 and Sample-F1 than the model trained without stopping.

## 3 Discussion

This is the first study to show that a DL model using only five lower limb IMUs can automatically annotate FOG episodes at the same granularity as clinical experts. We extended the MS-TCN model to enable sample-wise FOG annotations. Two clinical measures were computed to evaluate the FOG severity predicted by the DL model trained for the clinical setting (Model_Clinical), the %TF and #FOG (23). Model_Clinical showed a strong agreement with the experts’ observations for %TF (ICC=0.92) and #FOG (ICC=0.95). In previous studies, the ICC between independent raters on the TUG task was reported to be 0.87 (63) and 0.73 (23) for %TF and 0.63 (23) for #FOG, while for 360Turn, the ICC between raters was reported to be 0.99 for %TF and 0.86 for #FOG (20). While the ICC value in previous studies varied depending on the specific tool and population being studied (64), in comparison, our proposed model achieved similar levels of agreement. This holds significant promise for future AI-assisted FOG annotation work, whereby the DL model annotates FOG episodes initially, and the clinical expert verifies/adjusts only where required. Despite the high inter-rater reliability with the experts, results showed that the model statistically overestimated FOG severity with a higher %TF and #FOG than the experts when evaluating all trials. The overestimation of %TF and #FOG was partly due to false positives when predicting FOG-related movement, such as shuffling and festination, as FOG segments. Given that these FOG-related movements often lie on the boundary between freezing and non-freezing (43), it can be challenging for the model to accurately annotate and categorize them in a manner consistent with nearby FOG episodes.

Moreover, this study evaluated the generalization of DL models across tasks and medication states by comparing DL models trained for a specific task or medication state. Two technical metrics, Sample-F1 and Segment-F1@50, were computed and statistically assessed. Results showed no significant difference between the model trained across different FOG-provoking tasks and medication states (Model_Clinical) and the model trained only on a specific condition (Model_TUG, Model_360Turn, Model_Off, and Model_On). Next, we evaluated whether the DL model trained only on one FOG-provoking task or medication state could generalize to other FOG-provoking tasks or medication states. Results showed that the model trained on one FOG-provoking task (i.e., TUG or 360Turn) could better detect FOG in such a task than the model without training on such tasks. Although previous studies have shown that gait patterns are altered post anti-Parkinsonian medication (40; 41), our results also showed that the model trained on one medication state could still detect FOG in the other medication state. As a result, we recommend caution when applying DL-based FOG assessment models on FOG-provoking tasks that were not explicitly trained on, while applying models trained on different medication states does not show such discrepancies. This also has positive implications for future work toward daily-life FOG detection. Training data needs to be diversified for all activities encountered during daily life, especially during which FOG may be more presented. On the other hand, diversifying training data towards the medication states is unnecessary, making it more feasible as data can be collected in the On-medication intake regimens in the future.

While existing approaches utilized DL models to detect FOG on standardized FOG-provoking tasks with IMUs (33; 24), the model’s ability to distinguish FOG from stopping remains undetermined, which is critical for free-living assessment (43). Therefore, voluntary and instructed stops were introduced in the standardized FOG-provoking tasks. When evaluating trials without stops, results showed no difference between the model trained without stops and the model trained with stops, showing that adding stopping periods in the training data does not affect the DL model to detect FOG. Additionally, when evaluating trials with stops, results showed that compared with the model trained without stops, the model trained with stops produced less FP of stopping (16 compared to 74). While it was considered that stops are difficult to distinguish from FOG with movement-related signals, especially for akinetic FOG (65), our model could still detect FOG in the presence of stops. Moreover, our result highlights the importance of including stopping in the training data.

Although this study has provided valuable insights, there are some limitations to acknowledge. The first limitation is that the videos in our dataset were annotated sequentially by two clinical experts. The first annotator’s work was verified and, if needed, corrected by the second annotator. As a result, we could not calculate the inter-rater agreement in our study to compare our models’ annotation against. However, the literature shows that inter-rater agreement is 0.39%-0.99% (63; 23; 20; 27; 31; 24) and that these differences between experts were sometimes due to minor differences between FOG and FOG-related movements. Our DL model’s agreement with the clinical experts exceeded those previously published inter-rater agreements, and just as between experts, most of our model’s mispredicted FOG segments were marked as FOG-related segments by the experts. Future work could investigate the development of DL models that can better differentiate between FOG and FOG-related events. The second limitation is that this study simulated free-living situations by asking patients to stop when performing standardized FOG-provoking tasks. Yet, free-living movement will contain substantially more variance (e.g., daily activities) than captured during our standardized tasks. Moreover, FOG severity during our tasks does not necessarily represent FOG severity in daily life (42; 66). Therefore, future work should establish the reliability of our approach to data measured in free-living situations. The third limitation is that this study showed that training DL models with trials that include stopping resulted in better performance in detecting FOG in trials that include stopping. However, whether DL models are able to distinguish between FOG and stopping for all manifestations of FOG (e.g., akinetic FOG) remains to be investigated. The fourth limitation is the small number of participants compared to the other use cases in DL literature. As this study evaluated the model with the LOSO cross-validation approach, the results still showed that the model could generalize learned features to unseen subjects. Moreover, despite the small number of subjects, the number of samples and FOG events in the dataset used in this study is comparable with the literature (27; 33). Future studies could evaluate automatic FOG assessment on larger datasets or across datasets. The fifth limitation is that the recruited PD patients subjectively reported having at least one FOG episode per day with a minimum duration of five seconds. While the proposed model works for these severe freezers, it still has to be verified whether the model also generalizes to mild freezers.

## 4 Conclusion

In this paper, we introduced a DL model for IMU-based FOG assessment trained across two FOG-provoking tasks, i.e., timed-up-and-go and 360-degree turning-in-place, both On- and Off-medication states, and trials containing stopping. The neural network architecture, termed MS-TCN, was evaluated by comparing it with the experts’ observations on two clinical and two technical metrics. We established that the proposed DL model resulted in strong agreement with experts’ annotations on the percentage of time-frozen and the number of FOG episodes. This phenomenon highlights that a single DL model can be trained to generalize over FOG-provoking tasks and medication states for FOG assessment in a clinical setting. Moreover, we established that DL models should include specific FOG-provoking task in the training data in order to be able to detect FOG in such a task, while this is not necessary for different medication states. Finally, we established that the proposed model can still detect FOG in trials that contain stopping. Though, only when stopping is included in the training data. This phenomenon is encouraging and enables future work to investigate FOG assessment during everyday life.

## Data Availability

Our neural network architecture was implemented in Pytorch version 1.2 by adopting the public code repositories of Farha and Gall [38] and Pavlo et al. [53]. The datasets analyzed during the current study are not publicly available due to restrictions on sharing subject health information.

https://arxiv.org/abs/1903.01945

https://arxiv.org/abs/1811.11742

## List of abbreviations

PD: Parkinson’s Disease
FOG: Freezing of Gait
FOGQ: Freezing of Gait Questionnaire
NFOG-Q: New Freezing of Gait Questionnaire
DL: Deep Learning
TUG: Timed-Up-and-Go
360Turn: 360-degree Turning-In-Place
%TF: Percentage Time Spent Frozen
#FOG: Number of FOG Episodes
IMU: Inertial Measurement Unit
MoCap: Motion-captured
TCN: Temporal Convolutional Network
MS-TCN: Multi-Stage Temporal Convolutional Neural Network
LOSO: Leave-One-Subject-Out
SD: Standard Deviation
TP: True Positive
TN: True Negative
FP: False positive
FN: False Negative
ICC: Intra-class Correlation Coefficient
CI: Confidence Interval
BTK: Biomechanical Toolkit.

## Declarations

### Ethics approval and consent to participate

The study was approved by the local ethics committee of the UZ/KU Leuven (S65059) and all subjects gave written informed consent.

### Consent for publication

Not applicable

### Availability of data and materials

The input set was imported and labeled using Python version 2.7.12 with Biomechanical Toolkit (BTK) version 0.3 (67). The MS-TCN architecture was implemented in Pytorch version 1.2 (68) by adopting the public code repositories of MS-TCN (34), and VideoPose3D (51). All models were trained on an NVIDIA GeForce RTX 2080 GPU using Python version 3.7.11. The datasets analyzed during the current study are not publicly available due to restrictions on sharing subject health information.

### Author’s contributions

Study design by PY, BF, PG, AN, PS, and BV. Data analysis by PY. Design and implementation of the neural network architecture by PY, BF. Statistics by PY and BV. Subject recruitment, data collection, and data preparation by PG, MG1, MG2, and AN. The first draft of the manuscript was written by PY and all authors commented on subsequent revisions. The final manuscript was read and approved by all authors.

## Acknowledgements

We thank the participants for their willingness to participate.

